# Intraoperative Hydromorphone Decreases Post-Operative Pain Who Would Have Thought? An Instrumental Variable Analysis

**DOI:** 10.1101/2021.10.18.21263855

**Authors:** Brent Ershoff

## Abstract

**Background:** A growing body of literature suggests that intraoperative opioid administration can lead to both increased post-operative pain and opioid requirements. However, there has been minimal data regarding the effects of the intraoperative administration of intermediate duration opioids such as hydromorphone on post-operative outcomes. Causal inference using observational studies is often hampered by unmeasured confounding, where classical adjustment techniques, such as multivariable regression, are insufficient. Instrumental variable analysis is able to generate unbiased causal effect estimates in the presence of unmeasured confounding, assuming a valid instrumental variable can be found. We previously demonstrated, using a natural experiment, how hydromorphone presentation dose, i.e. the unit dose provided to the clinician, affects intraoperative administration dose, with the switch from a 2-mg to a 1-mg vial associated with decreased administration. As the change in hydromorphone presentation dose was unrelated to any external factors, presentation dose could serve as an instrumental variable to estimate the effect of intraoperative hydromorphone administration dose on post-operative outcomes.

**Methods:** In this observational study with 6,751 patients, an instrumental variable analysis was employed to estimate the causal effect of an increased intraoperative administration dose of hydromorphone on post-operative pain and opioid administration. The study population included patients who received intraoperative hydromorphone as part of an anesthetic at the University of California, Los Angeles, from October 2016 to November 2018. Before July 2017, hydromorphone was available as a 2-mg unit dose. From July 1, 2017 to November 20, 2017, hydromorphone was only available in a 1-mg unit dose. A two-stage least squares regression analysis was performed to estimate the effect of intraoperative hydromorphone administration dose on post-operative pain scores and opioid administration.

**Results:** An increase in hydromorphone administration caused a statistically significant decrease in Post-Anesthesia Care Unit pain scores as well as maximum and mean pain scores on post-operative days one and two, without a statistically significant effect on post-operative opioid administration. Various sensitivity analyses support the validity of the instrumental variable assumptions and suggest that the results are robust against violations of these assumptions.

**Conclusions:** The results of this study suggests that the intraoperative administration of intermediate duration opioids do not cause the same effects as short acting opioids with respect to post-operative pain. Instrumental variables, when identified, can be invaluable in estimating causal effects using observation data whereby unmeasured confounding is likely present.

## 1 Introduction

Perioperative administration of opioids is common and often necessary for the mitigation of intraoperative nociception and the treatment of post-operative pain.[1–3] Given their multitude of adverse effects, including sedation, respiratory depression, ileus, and postoperative nausea and vomiting (PONV), [4–6], many have argued for limiting or even abolishing the routine administration of intraoperative opioids, with several ERAS protocols calling for opioid-free anesthesia. [7–12]. In light of the international opioid epidemic, understanding the effects of perioperative opioid administration on post-operative pain as well as opioid consumption is paramount.

While opioids are the most efficacious treatment for the management of pain, intraoperative opioid administration has, paradoxically, been associated with increased post-operative pain and opioid requirements. There is substantial evidence suggesting that intraoperative opioid use, particularly the administration of ultra-short acting agents such as remifentanil, may contribute to hyperalgesia, with a subsequent increase in opioid requirements and pain in the post-operative period, [13] [14] with one meta-analysis indicating that patients receiving higher intraoperative opioid doses had higher pain scores than controls. The majority of these studies, however, failed to investigate the effect of the intraoperative administration of longer-acting opioids on post-operative outcomes. A recent randomized control trials (RCT) demonstrated that intraoperative methadone administration is associated with decreased post-operative pain[15]. However, these studies were limited to small sample sizes and were performed in rather uniform patient populations. To date, there is limited evidence as to whether increased intraoperative doses of hydromorphone, an intermediate duration opioid ,is associated with decreased post-operative pain, with a recent observational study suggesting that an increased dose was associated with higher post-operative pain scores [16]. As acute post-operative pain is a major problem that is inadequately treated and can have multiple significant adverse consequences, understanding the causal effect of intraoperative opioid administration on post-operative pain and opioid consumption is necessary for the rational use of these drugs. The goal of this study is to evaluate whether increasing the dose of intraoperative hydromorphone administration affects post-operative pain scores and opioid consumption.

While RCTs, when executed properly, are capable of estimating the *causal* effect of an intervention on an outcome, they are often limited by their excessive cost as well as the ability to generalize to other populations due to their strict inclusion and exclusion criteria. In observational studies, causal inference is challenged by the lack of random exposure assignment, thus hampering the ability to draw conclusions regarding causal effects. [17] As such, there exists the potential for either unmeasured or unknown confounders, i.e. variables that effect the outcome and differ in distribution across exposure status, which can bias the estimate of the casual effect of an exposure on the outcome. The Direct Acyclic Graph (DAG) [18] in Figure 3(a) illustrates the causal structure of an exposure *X* on an outcome *Y* with a set of confounders *U*. For instance, suppose *X* were defined as intraoperative hydromorphone dose, *Y* as post-operative pain score, and *U* as the type of surgical procedure. Performing simple linear regression, whereby *Y* is regressed on *X*, would yield biased estimates of the effect of *X* on *Y* as there exists a backdoor path from *X* to *Y*. The impact of measured confounders on the estimation of a causal treatment effect can be mitigated via a myriad of methods including propensity scores, regression, and matching [19, 20]. These methods, however, can only control for measured confounders and cannot account for the effect of unknown confounders.

Instrumental variable analysis is a causal inference technique that has the ability to estimate the causal effect of *X* on *Y* in observational studies even in the presences of unmeasured confounding. An instrument is defined as a variable that predicts the exposure, but conditional on the exposure, shows no independent association with the outcome.[21] Figure 3(b) While instrumental variable analyses are common in economics and other social science literature, they have been infrequently applied to solving causal inference problems in the medical sciences.[22] In this study, the effect of intraoperative hydromorphone administration on post-operative pain scores and opioid administration is examined using an Instrumental variable (IV) approach.

As we are interested in examining the effect of intraoperative hydromorphone administration on post-operative outcomes including pain scores and opioid requirements, a naive approach would be to regress these outcomes on intraoperative hydromorphone administration. This method, however, would undoubtedly yield biased estimates as there exists a host of confounders. For example, patients undergoing painful procedures are more likely to have both higher post-operative pain scores and have higher intraoperative opioid administration doses. Adjusting for known measured confounders can block this backdoor path, but there are other variables that are unknown to the investigator that cannot be controlled. For that reason, other techniques must be sought to more accurately estimate such causal effects.

Ershoff, et al [23] recently demonstrated that the presentation dose of hydromorphone, namely the unit dose in which it is available to clinicians, was associated with the quantity of hydromorphone administered in the intraoperative period. Using an interrupted time series analysis, the authors found that a decrease in the presentation dose of hydromorphone from 2mg to 1mg was associated with a significant decrease in the quantity of hydromorphone administered. Furthermore, when hydromorphone was reintroduced as a 2mg vial, the quantity administered increased. A major strength of this study was the fact that the switch in presentation dose was due to a change in pharmaceutical supply and was unrelated to any other policy change, and as such, was exogenous to the system. Thus, the distribution of covariates, including possible confounding variables, would not be expected to differ across the cohorts. In essence, this approximates a natural experiment whereby the treatment received, i.e. whether one was exposed to a 2mg or 1mg presentation dose, was randomized, but the researcher was not in control of the randomization. Therefore, as presentation dose affects the quantity of hydromorphone administered intraoperatively, and is unrelated to any other potential confounders, presentation dose may be a suitable instrumental variable for this study

The aims of this manuscript are two fold: first, to characterize the effect of an increased intraoperative administration dose of hydromorphone on post-operative pain scores and opioid requirements, and second, to demonstrate the use of an instrumental variable in answering causal inference questions in the presence of unmeasured confounding.

## 2 Materials and Methods

### 2.1 Data Extraction

This study qualified for Institutional Review Board (IRB) exception status by virtue of having no direct patient contact and using a deidentified dataset (IRB No. 15–000518). The data were obtained via the previously published perioperative data warehouse [24] which is a structured reporting schema that contains a vast amount of clinical data that were entered into the institution’s electronic medical record. Only patients who received hydromorphone during their anesthesia care, underwent surgery at one of University of California, Los Angeles (UCLA)’s two inpatient hospitals, and who had their recovery in the Post-Anesthesia Care Unit (PACU) were included in the primary analyses. In sensitivity analyses, data from patients who underwent surgery, but who did not receive hydromorphone as part of their anesthetic, were analyzed. The exposure variable, that is the total intraoperative hydromorphone dose administered, was calculated from the anesthesia start-time to the anesthesia end-time according to the anesthetic record.

### 2.2 Study Design

In this observational cohort study, we used an instrumental variable design to evaluate whether intraoperative hydromorphone administration causally affects various outcomes including post-operative pain scores and opioid administration over the first two days following surgery. The candidate instrumental variable is the presentation dose of hydromorphone, which is the unit dose of hydromorphone, in milligrams, contained in a single vial. As described in Ershoff, et al,[23] the dispensing of hydromorphone was performed electronically via Pyxis Anesthesia Medstations (CareFusion Corporation, USA) which were present in each anesthetizing location, and not at a central location. Pyxis Medstations use single-dose mini-drawer pockets for the management of controlled substances, where each pocket only stores, and thus dispenses, a single ampule of hydromorphone. The anesthesia provider could choose what percent of the unit dose to administer to the patient; additional hydromorphone beyond that contained in a unit dose could be administered by dispensing an additional unit dose. For example, if a provider dispensed a 2-mg vial and wished to administer 2.4 mg of hydromorphone, a second 2-mg vial would need to be dispensed, and 1.6 mg of hydromorphone would be returned at the end of the case.

This study sample included 6,751 adult patients who received intraoperative hydromorphone as part of an anesthetic during the study period. Patients less than 18 yr of age were excluded. Before July 1, 2017 (n = 4,186), hydromorphone was only dispensed to anesthesia providers in 2-mg vials (cohort 1). The dose of hydromorphone administered was at the discretion of the anesthesia provider. Any remaining hydromorphone was returned to pharmacy per our controlled substance reconciliation policy. On July 1, 2017, the 2-mg hydromorphone unit dose was removed from inventory and was replaced by 1-mg hydromorphone vials (cohort 2). The change in the hydromorphone unit dose from 2 mg to 1 mg was attributable to a change in the pharmaceutical supplier, and was unrelated to any other policy changes. This is crucial for the validity of the IV exchangeability assumption (see Instrumental Variable Assumptions). From July 1, 2017 to November 20, 2017 (n = 2,565), hydromorphone was only available in a 1-mg unit dose. In a similar manner, from November 21, 2017 onward, hydromorphone was reintroduced in the original 2-mg unit dose (cohort 3), with the 1-mg vial completely removed from inventory. The change from a 1-mg vial to back to a 2-mg vial, however, was associated with a nationwide shortage in hydromorphone, and therefore for this study, the study population excludes patients after November 20,2017. For the primary analysis, cohort 1 includes patients from 250 days prior to the change point and cohort 2 includes patients from the presentation dose switch to 142 days after the change point. As the time from the change point extends further back, the likelihood that the exchangeability assumption remains valid is decreased. Alternatively, as the time window around the change point is narrowed, the validity of the exchangeability condition becomes stronger, but the decreased number of subjects reduces statistical power. In sensitivity analyses, results are provided for study samples whereby cohort 1 is extended to include cases 500 days prior to the change point, or truncated to only 142 days prior to the change point. In additional analyses, subjects in cohort 3 (extending to 109 days after the second change point), representing the reintroduction of the 2-mg presentation dose, were included. For this analysis, cohorts 1 and 3 were combined into one group for presentation dose equal to 2mg, and cohort 2 represented its own group with presentation dose equal to 1mg.

#### Instrumental Variable Assumptions

In order for an IV analysis to provide unbiased estimates of the effect of an exposure on an outcome, several assumptions must be justified. Here, each of the assumptions of an IV analysis are discussed in the context of this study.

The DAG in Figure 1 illustrates a possible causal structure for *X, Y, U*, and *Z*. The goal is to estimate the causal effect of intraoperative hydromorphone dose on various outcome measures, as represented by the arrow from *X* to *Y*. The variable *U* represents the set of potential confounders for *X* and *Y*.

**Figure 1:**
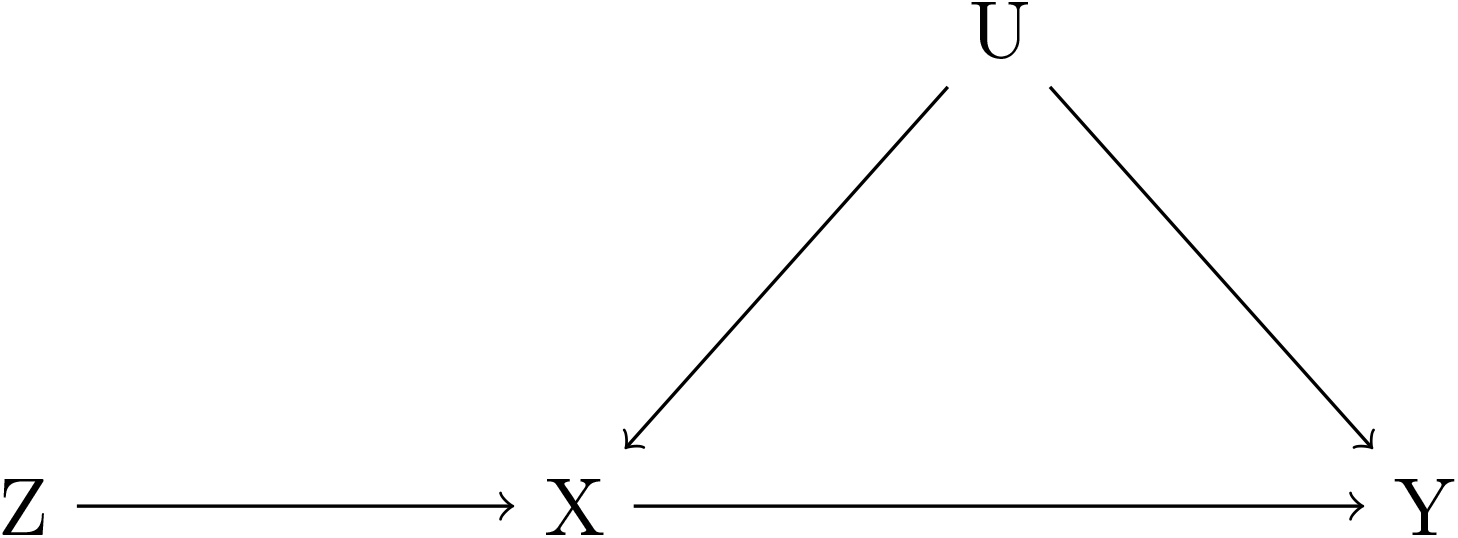
Direct Acyclic Graph of the Instrumental Variable Assumptions. *Z is an IV, X is the exposure, U is a set of confounders, and Y is an outcome. In the context of this study, Z represents the hydromorphone presentation dose, X represents the intraoperative hydromorphone administration dose, Y represents first PACU pain score, and U represents the set of confounders (e*.*g. age, sex, Body Mass Index (BMI)*,…*etc)*

**Figure 2:**
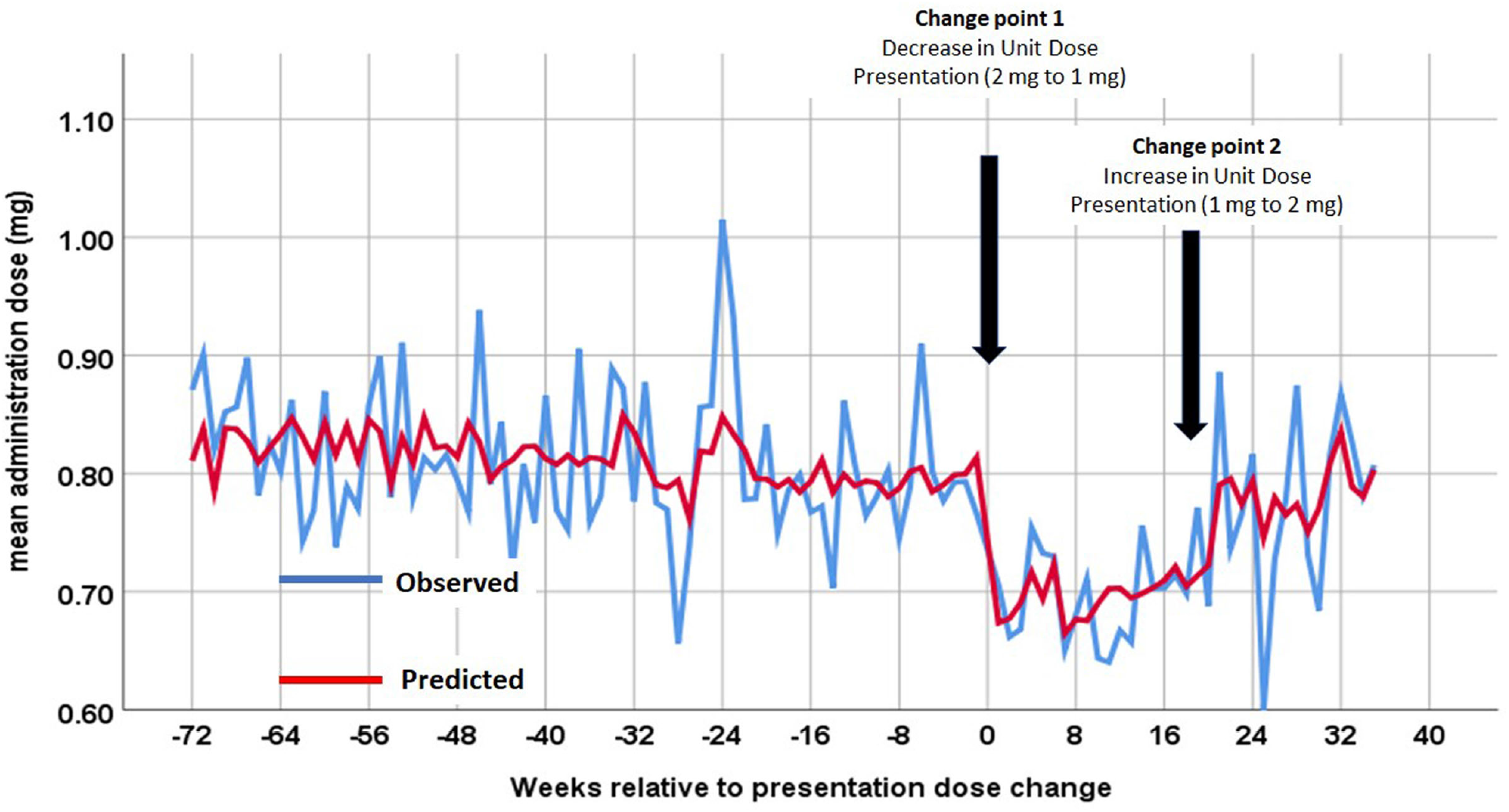
Interrupted Time Series Analysis: Hydromorphone Administration Dose Over Time. *A plot of the time series analysis illustrating the mean hydromorphone administration dose of hydromorphone, as a function of time (per week). The blue line indicates the observed mean administration dose within each week while the red line indicates the predicted mean administration dose within each week based on the segmented regression model. To generate the predicted value, individual predictions for each patient were computed using the multivariable segmented regression model and then aggregated at the weekly level. At the first change point, indicating the switch from a 2-mg to a 1-mg unit dose presentation, there was a statistically significant decrease in the mean hydromorphone administration dose. At the second change point (in week 20) indicating the switch from a 1 mg back to a 2-mg unit dose presentation, there was a significant increase in the mean administration dose of hydromorphone.Reprinted from [23] with permission*.

1. **The relevance assumption:** The relevance assumption states that the instrument *Z* is associated with the exposure *X*, that is 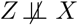. In this context, it states that hydromorphone presentation dose is associated with intraoperative hydromorphone administration dose. Of note, for the IV assumptions to hold, this association does not need to be causal [21], but in this case, Ershoff, et al provided strong evidence that indeed this relationship is causal. The relevance assumption of a *Z-X* association is one of the assumptions that can be empirically validated using data, and in fact, represents the first step of the Two-Stage Least Squares (TSLS) model described below. The strength of this association may be evaluated using F-statistics, and as a general rule, the instrument is declared weak if the F-statistic is less than 10 [17].
2. **The exclusion restriction**: *Z* affects the outcome *Y* only through *X*, that is *Z ╨ Y X, U*. This condition states that the association of the instrument *Z* on the outcome *Y* must be entirely mediated via the exposure. This is an assumption that cannot be proven empirically and must be supported by subject-matter knowledge to rule out the possibility of any direct effect of the instrument on the outcome [17, 25]. In the context of this study, such an assumption seems highly likely as it is difficult to imagine how the amount of hydromorphone contained within a vial could influence post-operative outcomes except through altering the quantity of hydromorphone administered to the patient. In a sensitivity analyses, the possibility of presentation dose directly affecting intraoperative fentanyl administration, which in turn, affects post-operative outcomes is consideredFigure 3(c). By conditioning on fentanyl administration, *Z* functions as a conditional instrumental variable and causal effect estimation of *X* on *Y* can still be drawn.[26]

**Figure 3:**
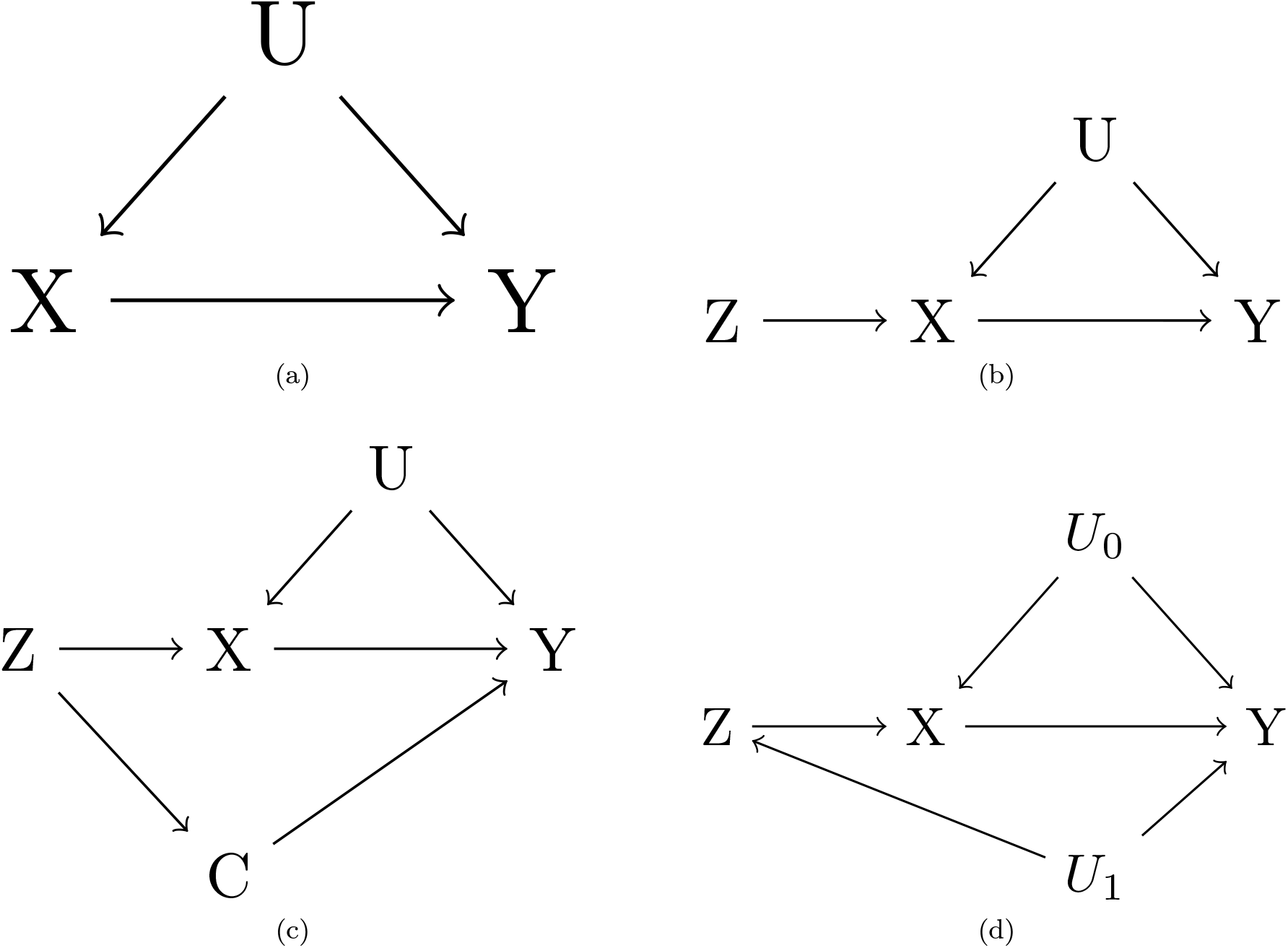
Direct Acyclic Graphs of Possible Causal Structures. *Direct Acyclic Graphs representing various causal structures. X is the exposure; Y is the outcome; U and U*_0_ *are the set of confounders of X and Y; Z is the instrumental variable; C is a mediator of the indirect path from Z to Y; U*_1_ *is the set of confounders for Z and Y*. ***a***: *U indicates the set of both measured and unmeasured confounders of X and Y. Regressing Y on X leads to biased causal effect estimates unless the backdoor path through U is blocked*. ***b***: *Z is an IV for the effect of X on Y*. ***c***: *Violation of exclusion restriction in the IV setting with variable C mediating the indirect path from Z to Y. Adjusting for C allows Z to be a conditional instrument of the effect of X on Y*. ***d***: *Violation of exogeneity assumption in the IV setting*.
3. **The exchangeability assumption:** *Z* does not share common causes with the outcome *Y* [17]. This assumption has also been termed the independence assumption [27], ignorable treatment assignment [28], or described as no confounding for the effect of *Z* on *Y* [25]. Therefore, the expected association between the set of possible confounders and the instrument is zero, that is *Z ╨ U*. As the switch in presentation dose was unrelated to any other policies or changes in the hospital system, the exposure assignment approximates random assignment. As such, there is no reason to expect that the patients who underwent an anesthetic in the group that was exposed to the presentation dose of 1mg were different from those exposed to a presentation dose of 2mg. The exchangeability assumption is partially verifiable in the data using measured covariates [17, 27]. Examining the distribution of covariates across these groups (Table 1) can support this conjecture, but it is not possible to empirically prove this.

**Table 1:** A Comparison of Covariates by Presentation Dose.
4. **Monotonicity**: The IV assumptions described above allow for identification of an upper and lower bound of the causal effect [17, 27, 29]. These bounds, however, are often wide and can be compatible with a positive, negative, or no effect. With the addition of a fourth assumption, namely monotonicity, point estimates can be calculated. [30] The monotonicity assumption states that the exposure is a non-decreasing function (or a non-increasing function) of the IV for all persons. In this context, it states that the increase in presentation dose from 1mg to 2mg led clinicians to either increase the quantity of hydromorphone administered intraoperatively, or to make no change at all. In observational studies, validation requires subject-matter knowledge and is difficult to test empirically.

|  | 2mg | 1mg | p-value |
| --- | --- | --- | --- |
| Number of Patients | 4186 | 2565 |  |
| Weight(kg) (mean (SD)) | 79.10 (20.26) | 79.94 (20.61) | 0.102 |
| BMI (mean (SD)) | 27.43 (6.47) | 27.70 (6.53) | 0.100 |
| Duration(minutes) (mean (SD)) | 246.71 (126.30) | 249.69 (135.51) | 0.359 |
| Age (mean (SD)) | 53.10 (16.78) | 53.28 (16.60) | 0.666 |
| Fentanyl(mg) (mean (SD)) | 0.14 (0.11) | 0.14 (0.12) | 0.397 |
| Morphine(mg) (mean (SD)) | 6.17 (3.64) | 6.75 (5.12) | 0.804 |
| Anesthesia type = General (%) | 4105 (98.1) | 2503 (97.6) | 0.212 |
| Sex = Male (%) | 1943 (46.4) | 1234 (48.1) | 0.184 |
| ASA Score (%) |  |  | 0.525 |
| 1 | 331 (7.9) | 182 (7.2) |  |
| 2 | 1850 (44.4) | 1123 (44.2) |  |
| 3 | 1900 (45.6) | 1177 (46.3) |  |
| 4 | 90 (2.2) | 58 (2.3) |  |
| 5 | 0 (0.0) | 1 (0.0) |  |
| ASA(E) = Emergent (%) | 197 (4.7) | 120 (4.7) | 1.000 |
| Ketamine = Yes (%) | 277 (6.6) | 148 (5.8) | 0.180 |
| Surgery Type (%) |  |  | 0.416 |
| 0 | 16 (0.4) | 11 (0.4) |  |
| 1 | 4148 (99.1) | 2534 (98.8) |  |
| 2 | 22 (0.5) | 20 (0.8) |  |

*Descriptive statistics of demographic and clinical characteristics are provided for patients stratified by presentation dose cohort membership. The distribution of covariates did not differ across the presentation dose cohorts. This finding supports the exchangeability assumption of IVs. Fentanyl, morphine, and ketamine variables refer to intraoperative administration. The variable "Surgery Type" indicates the surgical subspecialty group to which the patient belonged (see Covariates for description).*

### 2.3 Outcomes

- **Pain scores:** Pain scores were assessed based on the Numeric Pain Rating Scale (NPRS) and were measured at multiple time points in the post-operative period. In the PACU, pain scores were measured on both admission and discharge. Additional pain score assessments were recorded after discharge from the PACU, and the maximum and mean pain scores on the first and second post-operative days were calculated.
- **Morphine Milligram Equivalents (MME) administration:** The quantity of opioid administration in the post-operative period in both oral and intravenous formulations was calculated and described in MME. The total MME given within the first 60 minutes and 120 minutes of admission to the PACU were measured. Additionally, the total quantity of MME administered over the first and second post-operative days was measured.

### 2.4 Covariates

Several clinical and demographic covariates were measured to ensure that there were no significant differences in measured variables across the two cohorts. The exchangeability assumption states that there are no common causes of the instrument and the outcome. Given the quasi-random nature of IV assignment, there would not be expected to be an association between the IV and the set of covariates.[31] Table 1 reports summary statistics of the covariates across the two instrumental variable groups (namely 1mg versus 2mg presentation doses) Preoperative variables included patient age, sex, weight, BMI, and ASA Physical Status Classification System (ASA) (Schaumburg, Illinois), with one variable indicating the numeric component and another variable indicating the presence of the emergency modifier. Intraoperative covariates included case duration (defined as the difference of anesthesia end time and start time), intraoperative fentanyl and morphine administration doses, and an indicator for whether ketamine was given intraoperatively.

Other covariates included the primary surgical sub-specialty performing the procedure, as well as the anesthetic type (i.e., general anesthesia versus other). Because surgical sub specialty contained 25 levels, rather than creating 24 new binary categorical variables, the 25 categories were collapsed into three broader categories. Table 1. Group 0 includes cardiac surgical procedures, group 2 includes procedure categories that would not be expected to have high opioid requirements (i.e., dentistry, hematology, pediatric transplant hepatology, ophthalmology, pediatric hematology, and radiology), and group 1 refers to all other surgical sub-specialties.

There were minimal missing data among the covariates with values for ASA missing for 39 (0.6%) of patients. The mechanism of missing data was assumed to be missing completely at random, and therefore a complete case analysis was performed which would not be expected to bias coefficient estimates. Erroneous values based on clinician judgment were removed, including one for weight (value of 0), two for body mass index (values of 0 and 2,914), three for hydromorphone (values of 50, 50, and 11.2), and one for case duration of 1,888 minutes.

### 2.5 Statistical Analysis

Patient characteristics and study variables were summarized across IV groups using means ± SD and frequencies (%) unless otherwise noted. Characteristics were formally compared across groups by using two sample t-test and the chi-square test for continuous and categorical outcomes, respectively. Linear regression was used to assess base models from which to compare the IV analyses. For IV regression, TSLS [17, 27] was performed using the R package ivreg.[32] TSLS is the most common IV estimate technique whereby the first stage predicts the expected value of the exposure based on the instrument in a linear model: *E*[*X* | *Z*] = *α*_0_ + *α*_1_*Z*. The second stage then predicts the outcome as a function of the predicted exposure from the first stage: *E*[*Y* | *Z*] = *β*_0_ + *β*_1_*E*[*X* | *Z*].[21] Here, the parameter *β*_1_ is equivalent to the IV estimator. Of note, any measured covariate to predict the exposure may be added in the first stage and again in the second stage. Conditioning on these covariates will relax the assumption of marginal exchangeability to an assumption of conditional exchangeability based on the covariates [27]. All statistical analysis were performed using R [33] with tables generated using the R packages tableone [34] and modelsummary [35]. DAGs were generated using DAGitty [36]

#### 2.5.1 Sensitivity Analysis

While the IV assumptions, when valid, allow for unbiased causal effect estimates, it is important to determine how the estimates may change in response to violations of these assumptions, and thereby bolster the validity of the estimates.

1. **Time window size:** As the time from the change point increases, it becomes possible that there may be changes in the composition of patients across the groups, which calls into question the validity of the exchangeability assumption. As such, for the primary analysis, a smaller time window, i.e. only 250 days prior to the change point, was used. In sensitivity analyses, the time window around the change point was increased to 500 days prior to the change point, as well as shrunken to only 142 days prior to the change point, to demonstrate how the choice of the time window width did not qualitatively alter the results. Furthermore, the time window was extended to an additional 110 days after the re-introduction of the 2mg presentation dose, to illustrate how inclusion of these additional patients did not qualitatively affect the results. (see Study Design).
2. **Unknown confounding:** To demonstrate robustness against minor violations of the exchangeability assumption, the degree of confounding that would need to be present such that the null hypothesis would not be rejected was evaluated using sensitivity analysis. [37]. The exchangibility assumption states that there can be no common causes of *Z* and *Y*. The DAG in Figure 3(d) illustrates the causal structure whereby there exists a set of confounders, *U*_1_ which opens a backdoor path between *Z* and *Y*. The target quantity of IV estimation consists of a ratio of two population regression coefficients:

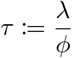

where *ϕ* and *λ* are the population regression coefficients of *Z* on *X* (the first stage) and *Z* on *Y* (the reduced stage), respectively. [38] If assessing the strength of confounders necessary to bring the IV point estimate to zero, or to not reject the null hypothesis of zero effect is all that is required, then the results of the sensitivity analysis of the reduced form is all that is needed. That is, if sensitivity analysis of the reduced form reveals that confounding is not strong enough to explain away *λ*, then it also cannot explain away the IV point estimate. The R package Sensemakr [37] was used to calculate how strong a potential confounder must be associated with both the exposure and the outcome in order for the reduced form to no longer be statistically significant, which in turn would cause the TSLS estimate to not be statistically significant.
3. **Falsification test:** A falsification test was performed whereby a control population that was *not* affected by the instrument (i.e. hydromorphone presentation dose) was shown to not experience an effect on the outcomes variables across the time periods associated with the presentation dose changes.[39]. Specifically, this population consisted of patients who did not receive hydromorphone intraoperatively, and therefore the change in presentation dose from 2mg to 1mg would have had no effect on intraoperative hydromorphone administration. If, however, there were other reasons for a change in outcome measures aside from that which was due to the change in intraoperative hydromorphone administration, one would expect that these changes would also affect this control population. Using linear regression, the association between presentation dose and the outcome measure among those in the control population were assessed. If a confounder, *U*_1_ existed as in Figure 3(d), one would expect to see a significant effect when regressing *Y* on *Z* in the control population.
4. **Exclusion restriction violation:** To evaluate the robustness of the exclusion restriction assumption, a casual structure whereby the presentation dose directly affected intraoperative fentanyl administration, which then affected the outcome variable was evaluated. While the possibility that the IV also directly led to clinicians changing their intraoperative fentanyl administration behavior cannot be entirely excluded, Figure 3(c) illustrates a conditional IV analysis where the causal effect of *X* on *Y* can still be estimated by blocking the path through *C*.

## 3 Results

### 3.1 Group Comparisons

Table 1 displays the summary statistics for relevant baseline covariates stratified by IV group. As expected, there were no statistically significant differences across the two groups which supports the exchangeability assumption of IVs.

### 3.2 Base Models

Simple linear regression yields biased estimates of the effect of intaoperative hydromorphone administration on post-operative pain and opioid administration. Such an analysis showed that an increase in intraoperative hydromorphone administration was associated with increased pain scores both in the PACU as well as post-operatively on days 1 and 2 Table 2. Patients who received higher doses of intraoperative hydromorphone received higher doses of opioids both in the PACU as well as post-operatively Table 2. Multivariable regression was performed whereby several covariates, some of which theoretically could function as confounders of the effect of *X* on *Y*, were included in these models. Table A1 and Table A2 display the effect estimates of intraoperative hydromorphone administration on the outcome measures based on a multivariable linear regression model. The effect estimates and p-values were qualitatively unchanged compared to the simple regression models.

**Table 2:** Simple Linear Regression Model

| Postoperative Pain Scores |  |  |  |  |  |  |
| --- | --- | --- | --- | --- | --- | --- |
|  | First PACU Pain | Last PACU Pain | Ave Pain (day 1) | Ave Pain (day 2) | Max Pain (day 1) | Max Pain (day 2) |
|  | Estimate= 0.25 | Estimate= 0.15 | Estimate= 0.14 | Estimate= 0.15 | Estimate= 0.15 | Estimate= 0.17 |
|  | CI=0.22, 0.28 | CI=0.12, 0.17 | CI=0.12, 0.16 | CI=0.13, 0.17 | CI=0.13, 0.18 | CI=0.14, 0.19 |
|  | p = <0.01 | p = <0.01 | p = <0.01 | p = <0.01 | p = <0.01 | p = <0.01 |
| Num.Obs. | 6505 | 6505 | 6662 | 5883 | 6662 | 5883 |

| Postoperative Opioid Administration |  |  |  |  |
| --- | --- | --- | --- | --- |
|  | MME PACU (60min) | MME PACU (120min) | MME (day1) | MME (day2) |
|  | Estimate= 1.47 | Estimate= 0.49 | Estimate= 5.14 | Estimate= 3.66 |
|  | CI=0.83, 2.10 | CI=0.42, 0.55 | CI=4.77, 5.51 | CI=2.81, 4.51 |
|  | p = <0.01 | p = <0.01 | p = <0.01 | p = <0.01 |
| Num.Obs. | 6595 | 6552 | 6663 | 5885 |

The table displays the effect estimates, 95 % confidence intervals, and p-values for simple linear regression models evaluating the association between intraoperative hydromorphone administration and various outcome measures. The effect estimates are based on a 0.2mg increase in intraoperative hydromorphone administration. CI = 95 % confidence interval. p= p-value. Ave Pain and Max Pain indicates the average and maximum pain scores over the indicated day, respectively. MME includes both oral and intravenous administration of opioids.

### 3.3 Instrumental Variable Analysis

Table 3 displays the effect size estimates, confidence intervals, and p-values for the effect of intraoperative hydromorphone administration on PACU and postoperative pain scores from the TSLS models. Intraoperative hydromorphone caused *decreased* pain scores at all time points including admission and discharge PACU pain scores, as well as mean and maximum pain scores on post-operative days one and two. Table 3 displays the estimates for the TSLS model with post-operative opioid administration doses as the outcome variables. For all time points examined, there were no statistically significant effects of intraoperative hydromorphone administration on MME administration. In additional analyses, whether the addition of measured covariates (Table 1) to the TSLS model would qualitatively change the main findings was explored. Table A3 and Table A4 display the effect estimates for an IV analysis which includes potential confounders and show that the results remain qualitatively unchanged. A diagnostic test for weak instruments, which is an F test of the first stage regression, was greater than 60 for all models suggesting presentation dose was not a weak instrument.

**Table 3:**
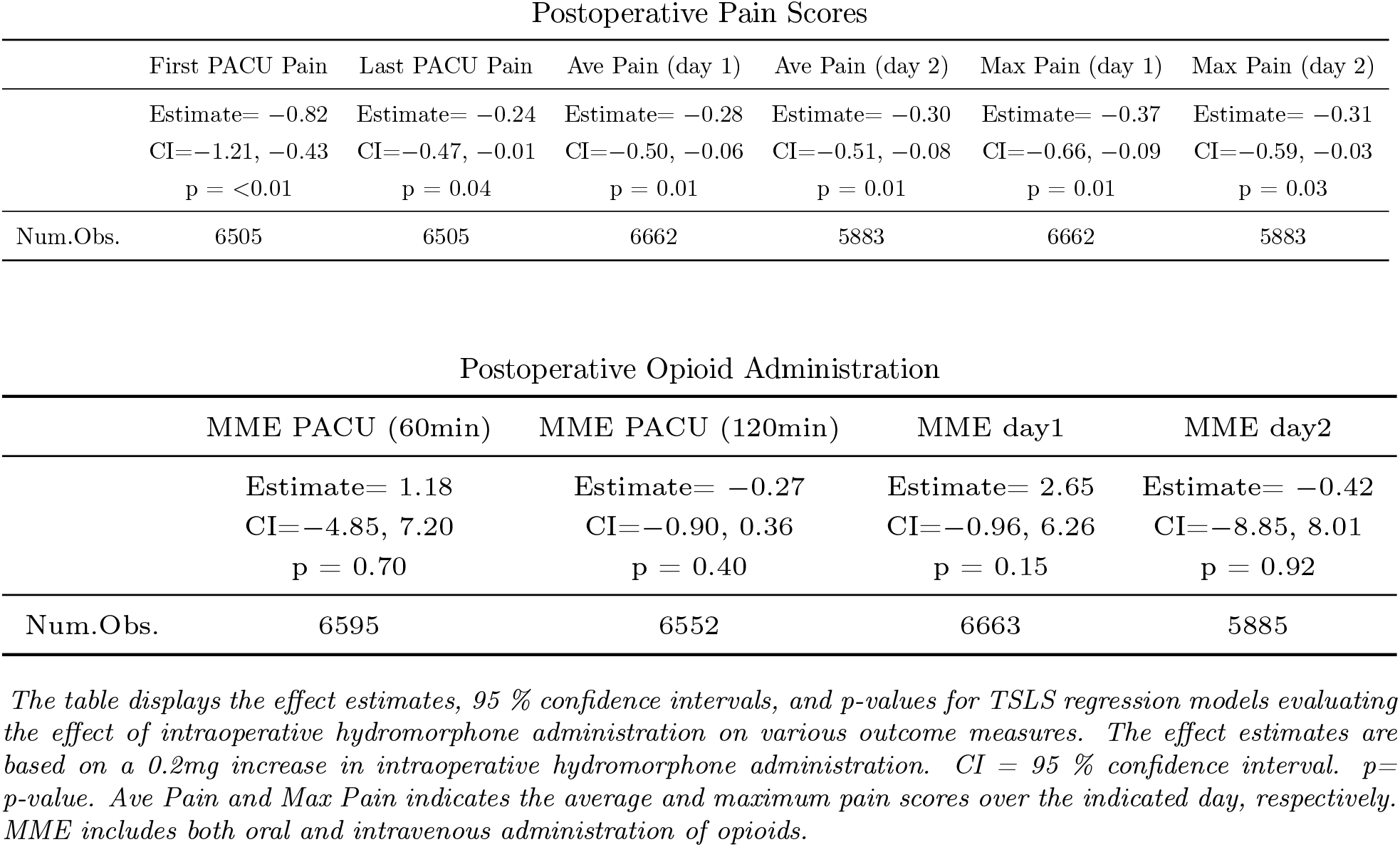
TSLS Regression Models

#### 3.3.1 Sensitivity Analysis Results

Several sensitivity analyses were performed to determine the robustness of the IV assumptions to different causal structures as well as how results would change in response to violations of the assumptions.

1. **Time window size** As the time window around the change point increases in size, the argument of exchangibility between the two presentation cohorts becomes less plausible. To provide further evidence in support of the findings, TSLS estimates for samples with varying time window sizes were calculated. Below, the TSLS are provided for a smaller time window where cohort 1 (i.e. the 2mg presentation dose cohort) extends only 142 days prior to the change point. The narrower time window increases the validity of the exchangeability assumption. The effect of intraoperative hydromorphone on all post-operative pain score measures were qualitatively unchanged, with increased intraoperative hydromorphone administration causing lower post-operative pain scores (Table 4). Results for a study population with a wider time windows extending 500 days prior to the change point, as well as a population with a time window extending forward in time to include the re-introduction of the 2mg presentation dose, are provided in Table A6 and Table A7. The results are qualitatively unchanged in all analyses.

**Table 4:** TSLS Regression with Narrow Time Window
2. **Unknown confounding** As IV estimation consists of a ratio of two population regression coefficients (equation 1), demonstrating that the reduced form, namely the *Z-Y* association, is statistically different from zero is all that is necessary in order to show that a potential confounder could not qualitatively alter the statistical significance of the TSLS estimate. For each of the pain score outcomes evaluated, the percentage of residual variation with both the exposure and the outcome that an unknown confounder would have to explain in order to bring the lower bound of the confidence interval to zero, was calculated. For example, for the outcome first PACU pain score, a potential confounder would have to explain 3.8 % of the residual variance of both the outcome as well as presentation dose, in order to explain away all the observed effect. For perspective, BMIand intraoperative fentanyl administration only accounts for 0.1 % and 0.4% of the variance of this outcome, respectively. For last PACU pain score, maximum pain scores on day 1 and 2, and mean pain score on days 1 and 2, the residual variance needed to explain away the observed effect is 2.5 %, 3.4%, 3.9%, 3.3%, and 3.0%, respectively. Figure 3 is a contour plot of the range of possible t-values that confounders with different strengths would cause. For first PACU pain score, confounders that are up to 40 times stronger than that of BMI would not change the statistical significance of the reduced form. Collectively, these results support that the IV estimates were not sensitive to violations of the exchangeability assumption.
3. **Falsification test** A control population of patients who underwent an anesthetic but did not receive intraoperative hydromorphone was examined. As discussed above, the reduced form estimate must be statistically significant in order for the TSLS estimate to be significant. Therefore, the reduced form estimates were compared for the population of patients who received hydromorphone and those who did not. If a set of confounding variables, *U*_1_, were actually responsible for the association between *Z* and *Y*, then one would expect that such a relationship would exist in the population not exposed to intraoperative hydromorphone. Table 5 shows the simple linear regression estimates for both of these populations. While there were statistically significant effects for each of the outcome measures in the population that received intraoperative hydromorphone, there were no such significant effects within the control population. These findings strengthen both the exchangeability and the exclusion assumptions. If there existed confounding between *Z* and *Y* as in Figure 3(d), one would expect to see a significant effect when *Y* was regressed on *Z* in the control population.

|  | First PACU Pain | Last PACU Pain | Ave Pain (day 1) | Ave Pain (day 2) | Max Pain (day 1) | Max Pain (day 2) |
| --- | --- | --- | --- | --- | --- | --- |
|  | Estimate= -0.90 | Estimate= -0.37 | Estimate= -0.32 | Estimate= -0.38 | Estimate= -0.44 | Estimate= -0.42 |
|  | CI=-1.42, -0.38 | CI=-0.69, -0.05 | CI=-0.61, -0.03 | CI=-0.67, -0.08 | CI=-0.83, -0.06 | CI=-0.81, -0.04 |
|  | p = <0.01 | p = 0.03 | p = 0.03 | p = 0.01 | p = 0.02 | p = 0.03 |
| Num.Obs. | 4784 | 4784 | 4907 | 4316 | 4907 | 4316 |

The table displays the effect estimates, 95 % confidence intervals, and p-values for TSLS regression models evaluating the effect of intraoperative hydromorphone administration on various outcome measures. In this sensitivity analysis, a narrow time window was utilized where the 2mg presentation dose cohort extends only 142 days prior to the change point. The effect estimates are based on a 0.2mg increase in intraoperative hydromorphone administration. CI = 95 % confidence interval. p= p-value. Ave Pain and Max Pain indicates the average and maximum pain scores over the indicated day, respectively. MME includes both oral and intravenous administration of opioids.

**Table 5:** Falsification Test
4. **Exclusion Restriction** If the exclusion restriction were violated by presentation dose directly affecting fentanyl administration, which in turn affected the outcome variable (Figure 3(c)), then conditioning on intraoperative fentanyl administration allows presentation dose to function as a conditional IV. Table A5 displays the effect estimates for the TSLS models with presentation dose as a conditional IV allowing for unbiased estimates of the effect of intraoperative hydromorphone on the outcome variables. The results are qualitatively unchanged from the primary analyses with increased intraoperative hydromorphone administration dose causing lower post-operative pain scores without a significant effect on post-operative opioid use.

| Population with Intraoperative Hydromorphone |  |  |  |  |
| --- | --- | --- | --- | --- |
|  | First PACU Pain | Last PACU Pain | Ave Pain (day 1) | Max Pain (day 1) |
|  | Estimate= 0.50 | Estimate= 0.15 | Estimate= 0.17 | Estimate= 0.23 |
|  | CI=0.31, 0.69 | CI=0.01, 0.28 | CI=0.05, 0.29 | CI=0.07, 0.38 |
|  | p = <0.01 | p = 0.03 | p = <0.01 | p = <0.01 |
| Num.Obs. | 6505 | 6505 | 6662 | 6662 |
| Population without Intraoperative Hydromorphone |  |  |  |  |
|  | First PACU Pain | Last PACU Pain | Ave Pain (day 1) | Max Pain (day 1) |
|  | Estimate= 0.06 | Estimate= -0.01 | Estimate= -0.06 | Estimate= -0.05 |
|  | CI=-0.08, 0.19 | CI=-0.11, 0.09 | CI=-0.15, 0.03 | CI=-0.20, 0.09 |
|  | p = 0.41 | p = 0.87 | p = 0.21 | p = 0.47 |
| Num.Obs. | 9664 | 9664 | 10 135 | 10 135 |

The table displays the effect estimates, 95 % confidence intervals, and p-values for linear regression models evaluating the association between intraoperative hydromorphone administration and various outcome measures for two distinct patient populations. The first sample consists of the primary patient population who received hydromorphone as part of the anesthetic whereas the control sample consists of patients who did not receive intraoperative hydromorphone. The linear regression model represents the reduced form of the IV [38] As expected, for the primary sample, the reduced form is statistically significant. Pain scores in the 1mg presentation cohort are higher on average than those in the 2mg cohort. However, for the control sample, the estimates for the reduced form are not statistically significant. These results bolster the exchangeability and exclusion assumption. If there were confounding or other paths from Z to Y, then one would expect a to observe statically significant effect in the control population. The effect estimates are based on a 0.2mg increase in intraoperative hydromorphone administration. CI = 95 % confidence interval. p= p-value. Ave Pain and Max Pain indicates the average and maximum pain scores over the indicated day, respectively.

## 4 Discussion

This large single center observational study demonstrates that an increased intraoperative administration dose of hydromorphone decreases post-operative pain scores without a concomitant increase in post-operative opioid administration. In contrast to most observational studies performed to date, which employ classical adjustment techniques such as multivariable regression or propensity score matching, this study used an instrumental variable approach to generate unbiased estimates of the effect of an exposure on the outcome in the presence of unmeasured confounding. Through the use of a natural experiment, whereby hydromorphone presentation dose was changed by a pharmaceutical supplier, the ability to study the causal effect of intraoperative hydromorphone administration on various outcomes could be interrogated. Importantly, the change in opioid supply was unrelated to any other policies within UCLA health, and therefore a patient who presented for an anesthetic either prior to the change point or after the change point could be considered “as if” randomly assigned. After carefully reviewing the assumptions of an IV analysis, and demonstrating how these assumptions are valid and robust against minor violations, the causal effect of increased intraoperative hydromorphone administration on post-operative pain is evident. While it may seem obvious that increasing the dose of intraoperative opioid administration, a class of drugs which has been known for decades to be a highly efficacious in the treatment of acute pain, leads to decreased pain, many recent studies have cast doubt on this assertion.

A growing body of research has suggested that the intraoperative administration of short and ultra-short acting opioids, including remifentanil and fentanyl, can produce a dose related increases in pain scores and opioid consumption in the post-operative period due to opioid induced hyperalgesia and tolerance. In contrast, methadone, which is both a long-acting opioid and an N-methyl-D-aspartate antagonist, has been shown to decrease pain in both the early post-operative period as well as after months of follow-up. To date though, these studies have been small in size and restricted to select populations. Timing of opioid administration has also been proposed as an important factor with some arguing that administration before the end of surgery has deleterious effects. [40]

Currently, there has been a dearth of data evaluating whether intraoperative hydromorphone, one of the most commonly administered intraoperative opioids, affects post-operative pain scores. Given its ubiquitous use, it is critical to understand hydromorphone’s effect in the control of post-operative pain and to avoid being misled by biased effect estimates from observational studies. Recently, Curry, et al performed an observational study evaluating the effect of hydromorphone dose on post-operative pain scores, and found that an increased intraoperative hydromorphone dose was associated with increased post-operative pain scores.[16] This study, however, attempted to adjust for potential confounders by using multivariable regression, which makes it susceptible to inadequate control of unmeasured confounding, likely biasing effect estimates. In this study, by using an instrumental variable analysis, unbiased causal effect estimates in the setting of unmeasured confounders can be calculated.

The adequate treatment of post-operative pain is of utmost importance for a number of reasons. Postoperative pain remains undertreated with up to 80 % of patients reporting inadequate post-operative pain control. This leads to a multitude of negative consequences including increased morbidity, impaired physical function, lower quality of life, slowed recovery, prolonged opioid use during and after hospitalization, and increased cost of care. [41–43] Post-operative pain is associated with postoperative cognitive decline and delirium,[44] surgical site infections,[45], hospital readmission.[46], and subjectively, a poorer recovery process, lower satisfaction, and more regret about having had surgery.[47] Inadequate control of acute pain is the greatest risk factor for the later development of chronic post-operative pain which affects up to 60 % of patients [48, 49], with severe pain at the surgical site on the day of surgery being a significant predictor of persistent opioid use 6 months post-operatively in patients undergoing total knee or hip arthroplasty.[50] Similarly, the severity of post-operative pain has been shown to be a key risk factor in the development of persistent post-operative pain (PPP), with adequate treatment of acute pain in the early post surgical period associated with a lower incidence of PPP.[51]

Aside from identifying the causal effect of hydromorphone on various outcome measures, this study is novel in that it uses an instrumental variable approach. The change in presentation dose via an exogenous event casts a unique opportunity to use this powerful statistical technique to address causal inference questions. Future studies can evaluate whether this effect holds in other populations and health centers. Furthermore, this study motivates the use of this technique to answer other questions where a valid instrument can be identified. As drug dose presentations are frequently changing, it is interesting to postulate what future studies can be surmised.

### 4.1 Limitations

While the IV analysis has several benefits over more common adjustment methods, this study was not a RCT, and deviations from the IV assumptions can bias effect estimates. While there were no reasons to believe that there existed other policies changes that occurred in proximity to the change in hydromorphone presentation dose, the possibility that such a change could have occurred, either altering the clinical and demographic characteristics of the study population, or altering how pain was managed, cannot be entirely excluded. Through various sensitivity analyses, including shrinking the time window around the change point and performing a falsification test, this threat of validity to the IV assumptions can be greatly mitigated, but not eliminated.

It is crucial to understand precisely what quantity the IV estimate is actually estimating. Under the monotonicity assumption, the IV estimate is actually a local average treatment effect, that is the effect of the exposure on the outcome among the subset of the populations that actually was affected by the instrument. Specifically, the IV analysis estimates the effect of an increased hydromorphone administration dose among the subset of patients who had their intraoperative hydromorphone administration dose increased in the context of the change in presentation dose. As such, one can not estimate the effect of an increased hydromorphone administration dose on the various outcomes among patients who would have received the same administration dose regardless of presentation dose.

As TSLS estimator is linear, if the exposure-outcome relationship is in fact, non-linear, the parameter estimate will approximate the population averaged causal effect, that is the average effect resulting from a uniform increase in the exposure for all persons in the population. [52] This approximation, however, will be poor if the change in exposure is much greater than that associated with the IV. For this analysis, the results are expressed in terms of a 0.2mg hydromorphone increase which is close to the average effect of the presentation dose change on the exposure. If linearity holds, then the effect of a .4mg increase in hydromorphone will simply be twice the effect of the 0.2mg. Of course, as the increase in hydromorphone gets larger, the linearity will no longer hold and clearly this study cannot extrapolate the effect of, for example, a 5mg increase in intraoperative hydromorphone administration.

### 4.2 Conclusion

The results from this large observational dataset provides evidence that increasing the dose of intraoperative hydromorphone, within the dose range observed in this study, decreased post-operative pain scores up until the second day, without a consequent increase in opioid administration. Given the plethora of negative consequences associated with post-operative pain, these findings should inspire further studies to elucidate these effects.

**Figure 3:**
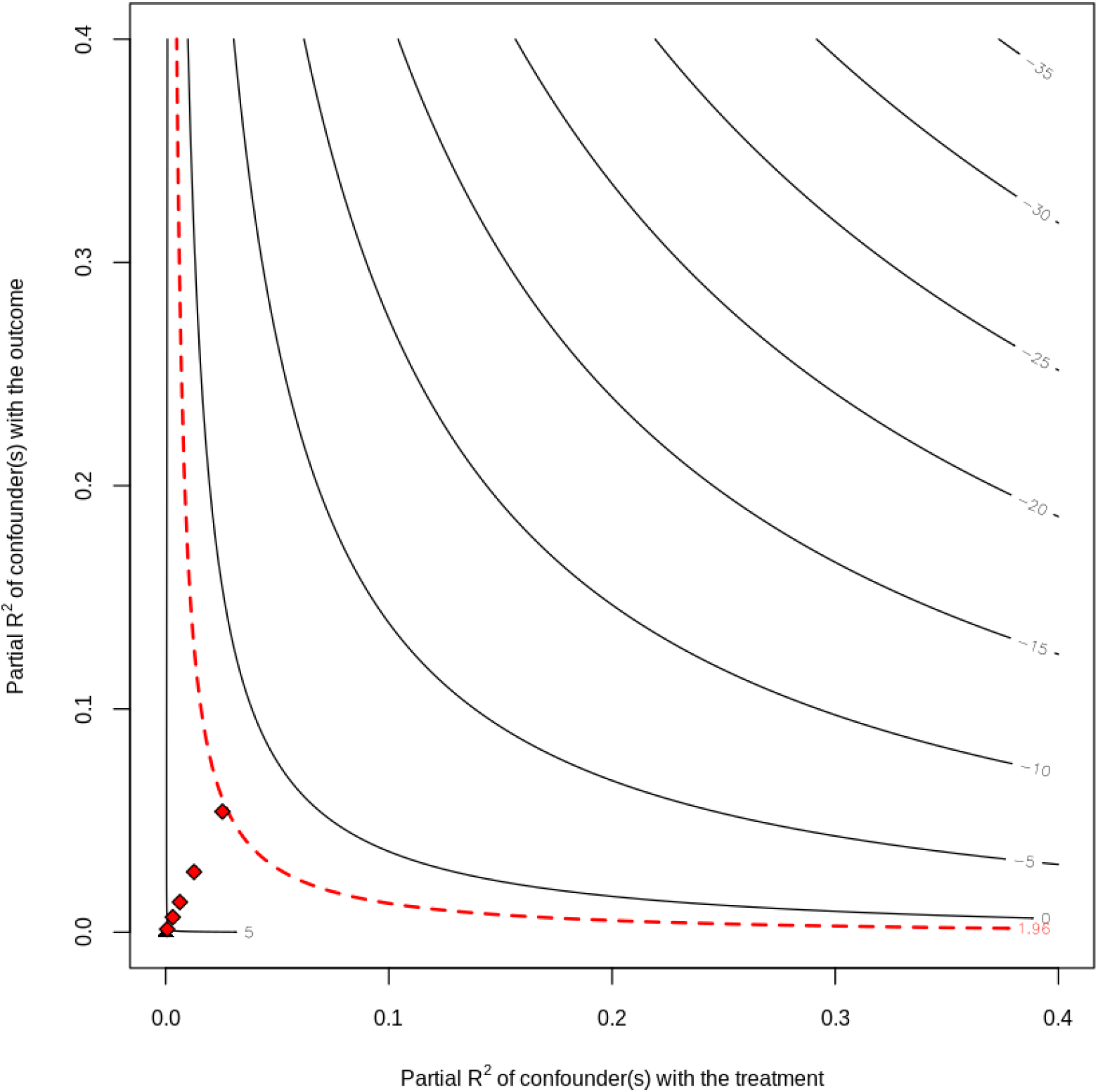
Sensitivity Contour Plot of t-values *Contour plot examining the sensitivity of the t-value for testing the null hypothesis of zero effect. The horizontal axis describes the fraction of the residual variation in the treatment (partial R*^2^*) explained by the confounder; the vertical axis describes the fraction of the residual variation in the outcome explained by the confounder. The contours show the adjusted estimate that would be obtained for an unobserved confounder (in the full model) with the hypothesized values of the sensitivity parameters (assuming the direction of the effects hurts the preferred hypothesis)[38]. The red points indicate confounding strengths that are 5,10,20, and 40 times as strong as BMI. The plot reveals that, at the 5% significance level, the null hypothesis of zero effect would still be rejected given confounders up to 40 times as strong as BMI, and thus the reduced form of the IV does not lose statistical significance. This suggests that the effect estimates are robust against confounding*

## Supporting information

STROBE Checklist

## Data Availability

The datasets generated during and/or analyzed during the current study are not publicly available due to institutional restrictions on data sharing and privacy concerns.

https://github.com/brentershoff/IV-Model

## 5 Appendix

**Table A1:**
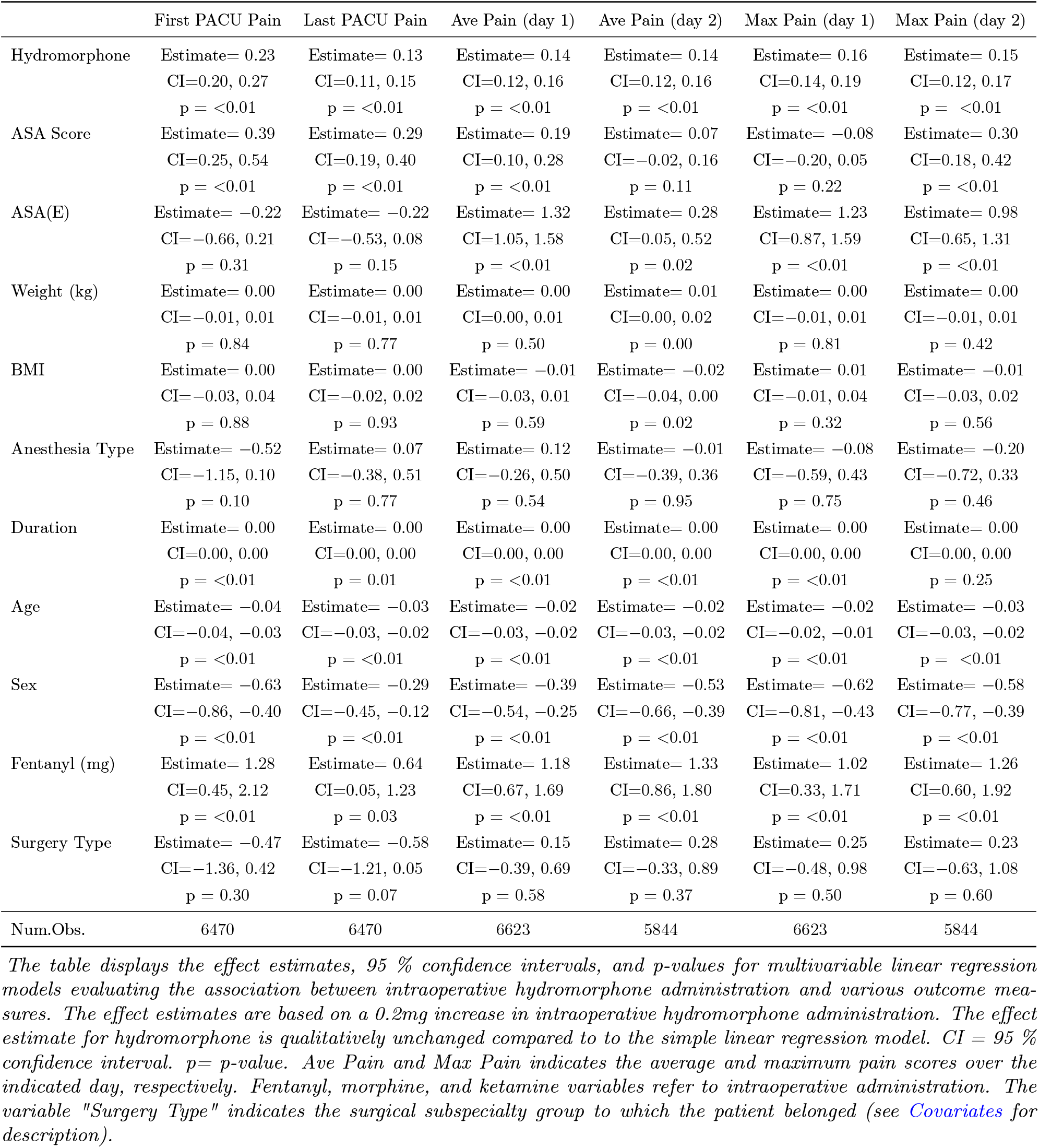
Multiple Regression Analysis: Pain Scores

**Table A2:** Multiple Regression Analysis: Opioid Administration

|  | MME PACU 60min | MME PACU 120min | MME day1 | MME day2 |
| --- | --- | --- | --- | --- |
| Hydromorphone | Estimate= 1.42<br>CI=0.75, 2.09<br>p = <0.01 | Estimate= 0.46<br>CI=0.39, 0.53<br>p = <0.01 | Estimate= 4.08<br>CI=3.74, 4.42<br>p = <0.01 | Estimate= 3.55<br>CI=2.67, 4.43<br>p = <0.01 |
| ASA Score | Estimate= -1.89<br>CI=-4.91, 1.14<br>p = 0.22 | Estimate= 0.31<br>CI=0.01, 0.61<br>p = 0.05 | Estimate= -4.61<br>CI=-6.17, -3.06<br>p = <0.01 | Estimate= 0.54<br>CI=-3.56, 4.64<br>p = 0.80 |
| ASA(E) | Estimate= -1.54<br>CI=-10.39, 7.31<br>p = 0.73 | Estimate= -0.52<br>CI=-1.40, 0.35<br>p = 0.24 | Estimate= 4.30<br>CI=-0.27, 8.86<br>p = 0.07 | Estimate= 11.63<br>CI=0.29, 22.97<br>p = 0.04 |
| Weight (kg) | Estimate= -0.11<br>CI=-0.33, 0.12<br>p = 0.35 | Estimate= 0.02<br>CI=0.00, 0.04<br>p = 0.12 | Estimate= 0.18<br>CI=0.07, 0.30<br>p = <0.01 | Estimate= 0.19<br>CI=-0.11, 0.50<br>p = 0.21 |
| BMI | Estimate= 0.14<br>CI=-0.54, 0.82<br>p = 0.68 | Estimate= -0.06<br>CI=-0.12, 0.01<br>p = 0.10 | Estimate= -0.24<br>CI=-0.59, 0.11<br>p = 0.18 | Estimate= -0.75<br>CI=-1.66, 0.16<br>p = 0.10 |
| Anesthesia Type | Estimate= -0.66<br>CI=-13.30, 11.99<br>p = 0.92 | Estimate= 0.32<br>CI=-0.94, 1.58<br>p = 0.62 | Estimate= 3.78<br>CI=-2.69, 10.26<br>p = 0.25 | Estimate= 2.44<br>CI=-15.38, 20.26<br>p = 0.79 |
| Duration | Estimate= -0.02<br>CI=-0.03, 0.00<br>p = 0.02 | Estimate= 0.00<br>CI=0.00, 0.00<br>p = 0.05 | Estimate= -0.02<br>CI=-0.03, -0.01<br>p = <0.01 | Estimate= -0.05<br>CI=-0.07, -0.03<br>p = <0.01 |
| Age | Estimate= -0.15<br>CI=-0.27, -0.03<br>p = 0.01 | Estimate= -0.02<br>CI=-0.03, -0.01<br>p = <0.01 | Estimate= -0.11<br>CI=-0.17, -0.05<br>p = <0.01 | Estimate= -0.44<br>CI=-0.60, -0.28<br>p = <0.01 |
| Sex | Estimate= 3.17<br>CI=-1.56, 7.91<br>p = 0.19 | Estimate= -0.09<br>CI=-0.56, 0.38<br>p = 0.70 | Estimate= -2.35<br>CI=-4.77, 0.06<br>p = 0.06 | Estimate= 2.17<br>CI=-4.24, 8.59<br>p = 0.51 |
| Fentanyl (mg) | Estimate= 21.80<br>CI=4.75, 38.85<br>p = 0.01 | Estimate= 3.53<br>CI=1.83, 5.23<br>p = <0.01 | Estimate= 175.34<br>CI=166.63, 184.06<br>p = <0.01 | Estimate= 42.43<br>CI=19.99, 64.86<br>p = <0.01 |
| Surgery Type | Estimate= -3.23<br>CI=-21.50, 15.04<br>p = 0.73 | Estimate= -1.08<br>CI=-2.89, 0.72<br>p = 0.24 | Estimate= -1.18<br>CI=-10.43, 8.08<br>p = 0.80 | Estimate= -6.63<br>CI=-35.70, 22.44<br>p = 0.65 |
| Num.Obs. | 6557 | 6516 | 6624 | 5846 |

The table displays the effect estimates, 95 % confidence intervals, and p-values for multivariable linear regression models evaluating the association between intraoperative hydromorphone administration and various outcome measures. The effect estimates are based on a 0.2mg increase in intraoperative hydromorphone administration. The effect estimate for hydromorphone is qualitatively unchanged compared to the simple linear regression model. CI = 95 % confidence interval. p = p-value. MME includes both oral and intravenous administration of opioids. Fentanyl, morphine, and ketamine variables refer to intraoperative administration. The variable "Surgery Type" indicates the surgical subspecialty group to which the patient belonged (see Covariates for description).

**Table A3:** IV Regression with Covariates: Pain Scores

|  | First PACU Pain | Last PACU Pain | Ave Pain (day 1) | Ave Pain (day 2) | Max Pain (day 1) | Max Pain (day 2) |
| --- | --- | --- | --- | --- | --- | --- |
| Hydromorphone | Estimate= -0.81 | Estimate= -0.23 | Estimate= -0.29 | Estimate= -0.28 | Estimate= -0.38 | Estimate= -0.27 |
|  | CI=-1.18, -0.44 | CI=-0.46, -0.01 | CI=-0.49, -0.08 | CI=-0.48, -0.09 | CI=-0.65, -0.10 | CI=-0.53, -0.02 |
|  | p = <0.01 | p = 0.04 | p = 0.01 | p = <0.01 | p = 0.01 | p = 0.04 |
| ASA Score | Estimate= 0.55 | Estimate= 0.35 | Estimate= 0.27 | Estimate= 0.13 | Estimate= 0.02 | Estimate= 0.36 |
|  | CI=0.36, 0.75 | CI=0.23, 0.47 | CI=0.16, 0.37 | CI=0.02, 0.23 | CI=-0.13, 0.16 | CI=0.22, 0.49 |
|  | p = <0.01 | p = <0.01 | p = <0.01 | p = 0.02 | p = 0.83 | p = <0.01 |
| ASA(E) | Estimate= -0.35 | Estimate= -0.27 | Estimate= 1.25 | Estimate= 0.21 | Estimate= 1.15 | Estimate= 0.91 |
|  | CI=-0.90, 0.19 | CI=-0.60, 0.06 | CI=0.95, 1.55 | CI=-0.07, 0.49 | CI=0.74, 1.55 | CI=0.54, 1.27 |
|  | p = 0.21 | p = 0.11 | p = <0.01 | p = 0.14 | p = <0.01 | p = <0.01 |
| Weight (kg) | Estimate= 0.01 | Estimate= 0.00 | Estimate= 0.01 | Estimate= 0.01 | Estimate= 0.01 | Estimate= 0.01 |
|  | CI=0.00, 0.02 | CI=0.00, 0.01 | CI=0.00, 0.01 | CI=0.01, 0.02 | CI=0.00, 0.02 | CI=0.00, 0.02 |
|  | p = 0.17 | p = 0.34 | p = 0.12 | p = <0.01 | p = 0.25 | p = 0.18 |
| BMI | Estimate= 0.00 | Estimate= 0.00 | Estimate= -0.01 | Estimate= -0.02 | Estimate= 0.01 | Estimate= -0.01 |
|  | CI=-0.04, 0.04 | CI=-0.02, 0.03 | CI=-0.03, 0.02 | CI=-0.04, 0.00 | CI=-0.02, 0.04 | CI=-0.04, 0.02 |
|  | p = 0.95 | p = 0.95 | p = 0.54 | p = 0.04 | p = 0.44 | p = 0.63 |
| Anesthesia Type | Estimate= -0.39 | Estimate= 0.11 | Estimate= 0.17 | Estimate= 0.14 | Estimate= -0.01 | Estimate= -0.04 |
|  | CI=-1.18, 0.40 | CI=-0.36, 0.58 | CI=-0.25, 0.60 | CI=-0.30, 0.58 | CI=-0.58, 0.55 | CI=-0.62, 0.53 |
|  | p = 0.33 | p = 0.64 | p = 0.43 | p = 0.53 | p = 0.96 | p = 0.89 |
| Duration | Estimate= 0.00 | Estimate= 0.00 | Estimate= 0.00 | Estimate= 0.00 | Estimate= 0.00 | Estimate= 0.00 |
|  | CI=0.00, 0.01 | CI=0.00, 0.00 | CI=0.00, 0.00 | CI=0.00, 0.00 | CI=0.00, 0.00 | CI=0.00, 0.00 |
|  | p = <0.01 | p = 0.11 | p = 0.87 | p = 0.15 | p = 0.89 | p = 0.02 |
| Age | Estimate= -0.06 | Estimate= -0.04 | Estimate= -0.03 | Estimate= -0.03 | Estimate= -0.03 | Estimate= -0.04 |
|  | CI=-0.07, -0.05 | CI=-0.04, -0.03 | CI=-0.04, -0.03 | CI=-0.04, -0.03 | CI=-0.04, -0.02 | CI=-0.05, -0.03 |
|  | p = <0.01 | p = <0.01 | p = <0.01 | p = <0.01 | p = <0.01 | p = <0.01 |
| Sex | Estimate= -0.61 | Estimate= -0.28 | Estimate= -0.40 | Estimate= -0.50 | Estimate= -0.63 | Estimate= -0.56 |
|  | CI=-0.91, -0.32 | CI=-0.46, -0.11 | CI=-0.56, -0.24 | CI=-0.66, -0.35 | CI=-0.84, -0.42 | CI=-0.76, -0.35 |
|  | p = <0.01 | p = <0.01 | p = <0.01 | p = <0.01 | p = <0.01 | p = <0.01 |
| Fentanyl (mg) | Estimate= 4.79 | Estimate= 1.86 | Estimate= 2.55 | Estimate= 2.66 | Estimate= 2.73 | Estimate= 2.60 |
|  | CI=3.16, 6.42 | CI=0.88, 2.84 | CI=1.69, 3.42 | CI=1.83, 3.48 | CI=1.58, 3.87 | CI=1.52, 3.68 |
|  | p = <0.01 | p = <0.01 | p = <0.01 | p = <0.01 | p = <0.01 | p = <0.01 |
| Surgery Type | Estimate= -0.95 | Estimate= -0.75 | Estimate= -0.05 | Estimate= 0.04 | Estimate= 0.00 | Estimate= -0.01 |
|  | CI=-2.09, 0.18 | CI=-1.43, -0.07 | CI=-0.67, 0.57 | CI=-0.67, 0.76 | CI=-0.82, 0.82 | CI=-0.95, 0.93 |
|  | p = 0.10 | p = 0.03 | p = 0.87 | p = 0.91 | p = 1.00 | p = 0.98 |
| Num.Obs. | 6470 | 6470 | 6623 | 5844 | 6623 | 5844 |

The table displays the effect estimates, 95 % confidence intervals, and p-values for TSLS regression model including additional covariates in evaluating the effect of intraoperative hydromorphone administration on various outcome measures. The effect estimate for hydromorphone is qualitatively unchanged compared to the TSLS model without covariates. The effect estimates are based on a 0.2mg increase in intraoperative hydromorphone administration. CI = 95 % confidence interval. p= p-value. Ave Pain and Max Pain indicates the average and maximum pain scores over the indicated day, respectively. Fentanyl, morphine, and ketamine variables refer to intraoperative administration. The variable "Surgery Type" indicates the surgical subspecialty group to which the patient belonged (see Covariates for description).

**Table A4:** IV Regression: Opioid Administration

|  | MME PACU 60min | MME PACU 120min | MME day1 | MME day2 |
| --- | --- | --- | --- | --- |
| Hydromorphone | Estimate= 0.96<br>CI=-4.93, 6.85<br>p = 0.75 | Estimate= -0.23<br>CI=-0.84, 0.38<br>p = 0.46 | Estimate= 2.04<br>CI=-1.09, 5.17<br>p = 0.20 | Estimate= -0.28<br>CI=-8.45, 7.89<br>p = 0.95 |
| ASA Score | Estimate= -1.81<br>CI=-5.00, 1.39<br>p = 0.27 | Estimate= 0.42<br>CI=0.09, 0.74<br>p = 0.01 | Estimate= -4.26<br>CI=-5.92, -2.61<br>p = <0.01 | Estimate= 1.05<br>CI=-3.21, 5.31<br>p = 0.63 |
| ASA(E) | Estimate= -1.60<br>CI=-10.48, 7.28<br>p = 0.72 | Estimate= -0.60<br>CI=-1.51, 0.30<br>p = 0.19 | Estimate= 3.98<br>CI=-0.66, 8.62<br>p = 0.09 | Estimate= 10.96<br>CI=-0.55, 22.46<br>p = 0.06 |
| Weight (kg) | Estimate= -0.10<br>CI=-0.33, 0.13<br>p = 0.38 | Estimate= 0.02<br>CI=0.00, 0.05<br>p = 0.05 | Estimate= 0.20<br>CI=0.08, 0.32<br>p = <0.01 | Estimate= 0.22<br>CI=-0.09, 0.53<br>p = 0.16 |
| BMI | Estimate= 0.14<br>CI=-0.54, 0.82<br>p = 0.68 | Estimate= -0.06<br>CI=-0.13, 0.01<br>p = 0.09 | Estimate= -0.24<br>CI=-0.60, 0.11<br>p = 0.18 | Estimate= -0.74<br>CI=-1.66, 0.17<br>p = 0.11 |
| Anesthesia Type | Estimate= -0.60<br>CI=-13.26, 12.07<br>p = 0.93 | Estimate= 0.40<br>CI=-0.90, 1.70<br>p = 0.55 | Estimate= 4.05<br>CI=-2.51, 10.60<br>p = 0.23 | Estimate= 3.86<br>CI=-14.32, 22.04<br>p = 0.68 |
| Duration | Estimate= -0.02<br>CI=-0.05, 0.01<br>p = 0.30 | Estimate= 0.00<br>CI=0.00, 0.00<br>p = 0.33 | Estimate= -0.01<br>CI=-0.03, 0.00<br>p = 0.17 | Estimate= -0.03<br>CI=-0.07, 0.01<br>p = 0.14 |
| Age | Estimate= -0.16<br>CI=-0.34, 0.01<br>p = 0.07 | Estimate= -0.03<br>CI=-0.05, -0.02<br>p = <0.01 | Estimate= -0.15<br>CI=-0.24, -0.06<br>p = <0.01 | Estimate= -0.52<br>CI=-0.76, -0.28<br>p = <0.01 |
| Sex | Estimate= 3.18<br>CI=-1.56, 7.91<br>p = 0.19 | Estimate= -0.08<br>CI=-0.56, 0.41<br>p = 0.75 | Estimate= -2.38<br>CI=-4.82, 0.06<br>p = 0.06 | Estimate= 2.37<br>CI=-4.10, 8.84<br>p = 0.47 |
| Fentanyl (mg) | Estimate= 23.30<br>CI=-2.35, 48.96<br>p = 0.07 | Estimate= 5.78<br>CI=3.13, 8.43<br>p = <0.01 | Estimate= 181.82<br>CI=168.59, 195.06<br>p = <0.01 | Estimate= 54.60<br>CI=20.33, 88.87<br>p = <0.01 |
| Surgery Type | Estimate= -3.44<br>CI=-21.92, 15.03<br>p = 0.71 | Estimate= -1.41<br>CI=-3.29, 0.48<br>p = 0.14 | Estimate= -2.13<br>CI=-11.59, 7.34<br>p = 0.66 | Estimate= -8.82<br>CI=-38.43, 20.80<br>p = 0.56 |
| Num.Obs. | 6557 | 6516 | 6624 | 5846 |

The table displays the effect estimates, 95 % confidence intervals, and p-values for TSLS regression model including additional covariates in evaluating the effect of intraoperative hydromorphone administration on various outcome measures. The effect estimate for hydromorphone is qualitatively unchanged compared to the TSLS model without covariates. The effect estimates are based on a 0.2mg increase in intraoperative hydromorphone administration. CI = 95 % confidence interval. p= p-value. MME includes both oral and intravenous administration of opioids. Fentanyl, morphine, and ketamine variables refer to intraoperative administration. The variable "Surgery Type" indicates the surgical subspecialty group to which the patient belonged (see Covariates for description).

**Table A5:** IV Regression Conditioning on Fentanyl

|  | First PACU Pain | Last PACU Pain | Ave Pain (day 1) | Ave Pain (day 2) | Max Pain (day 1) | Max Pain (day 2) |
| --- | --- | --- | --- | --- | --- | --- |
| Hydromorphone(mg) | Estimate= -0.83<br>CI=-1.22, -0.44<br>p = <0.01 | Estimate= -0.25<br>CI=-0.48, -0.01<br>p = 0.04 | Estimate= -0.29<br>CI=-0.51, -0.07<br>p = 0.01 | Estimate= -0.30<br>CI=-0.52, -0.09<br>p = 0.01 | Estimate= -0.38<br>CI=-0.67, -0.09<br>p = 0.01 | Estimate= -0.32<br>CI=-0.60, -0.04<br>p = 0.02 |
| Fentanyl(mg) | Estimate= 6.10<br>CI=4.01, 8.18<br>p = <0.01 | Estimate= 2.46<br>CI=1.21, 3.71<br>p = <0.01 | Estimate= 2.72<br>CI=1.62, 3.83<br>p = <0.01 | Estimate= 3.15<br>CI=2.08, 4.22<br>p = <0.01 | Estimate= 2.99<br>CI=1.52, 4.45<br>p = <0.01 | Estimate= 3.22<br>CI=1.82, 4.62<br>p = <0.01 |
| Num.Obs. | 6505 | 6505 | 6662 | 5883 | 6662 | 5883 |

The table displays the effect estimates, 95 % confidence intervals, and p-values for TSLS regression model including intraoperative fentanyl as a covariate. By adjusting for fentanyl, presentation dose functions as a conditional IV and unbiased estimates of the effect of intraoperative hydromorphone administration on the outcome can be generated. 3(c). The effect estimate for hydromorphone is qualitatively unchanged compared to the baseline TSLS model. The effect estimates are based on a 0.2mg increase in intraoperative hydromorphone administration. CI = 95 % confidence interval. p= p-value.

**Table A6:** TSLS Regression with Wider Time Window

|  | First PACU Pain | Last PACU Pain | Ave Pain (day 1) | Ave Pain (day 2) | Max Pain (day 1) | Max Pain (day 2) |
| --- | --- | --- | --- | --- | --- | --- |
|  | Estimate= -0.90<br>CI=-1.25, -0.55<br>p = <0.01 | Estimate= -0.30<br>CI=-0.50, -0.09<br>p = <0.01 | Estimate= -0.34<br>CI=-0.53, -0.15<br>p = <0.01 | Estimate= -0.26<br>CI=-0.44, -0.09<br>p = <0.01 | Estimate= -0.44<br>CI=-0.69, -0.19<br>p = <0.01 | Estimate= -0.33<br>CI=-0.57, -0.09<br>p = 0.01 |
| Num.Obs. | 10 572 | 10 572 | 10 857 | 9601 | 10 857 | 9601 |

The table displays the effect estimates, 95 % confidence intervals, and p-values for TSLS regression model with a wider time window that extends 500 days prior the change in presentation dose from 2mg to 1mg. The effect estimate for hydromorphone is qualitatively unchanged compared to the baseline TSLS model extending only 250 days prior to the change point. The effect estimates are based on a 0.2mg increase in intraoperative hydromorphone administration. CI = 95 % confidence interval. p= p-value. Ave Pain and Max Pain indicates the average and maximum pain scores over the indicated day, respectively.

**Table A7:** TSLS Regression with Extended Time Window

|  | First PACU Pain | Last PACU Pain | Ave Pain (day 1) | Ave Pain (day 2) | Max Pain (day 1) | Max Pain (day 2) |
| --- | --- | --- | --- | --- | --- | --- |
|  | Estimate= -0.73<br>CI=-1.11, -0.34<br>p = <0.01 | Estimate= -0.27<br>CI=-0.51, -0.03<br>p = 0.03 | Estimate= -0.30<br>CI=-0.52, -0.09<br>p = 0.01 | Estimate= -0.27<br>CI=-0.48, -0.07<br>p = 0.01 | Estimate= -0.46<br>CI=-0.76, -0.17<br>p = <0.01 | Estimate= -0.31<br>CI=-0.58, -0.04<br>p = 0.02 |
| Num.Obs. | 7541 | 7541 | 7741 | 6832 | 7741 | 6832 |

The table displays the effect estimates, 95 % confidence intervals, and p-values for TSLS regression model with a time window that extends 109 days past the reintroduction of the 2-mg presentation dose. The effect estimate for hydromorphone is qualitatively unchanged compared to the baseline TSLS model that does not include cohort 3 Study Design. The effect estimates are based on a 0.2mg increase in intraoperative hydromorphone administration. CI = 95 % confidence interval. p= p-value. Ave Pain and Max Pain indicates the average and maximum pain scores over the indicated day, respectively.

